# Rural-Urban Differences in Hospitalization Outcomes Among Young Adults (18-45) With Heart Failure, 2016-2022

**DOI:** 10.64898/2026.08.21.26361079

**Authors:** Heather R. Sherr, Wacim Benyoucef, R.J. Waken, Karen E. Joynt Maddox, Erin D. Solomon, Vi-Anh Hoang, Gmerice Hammond

**Affiliations:** Division of Cardiology, School of Medicine, Washington University, St. Louis, Missouri, USA; School of Medicine, University of Missouri, Columbia, Missouri, USA; Institute of Informatics, Data Science, and Biostatistics, School of Medicine, Washington University, St. Louis, Missouri, USA; Center for Advancing Health Services, Policy and Economics Research, School of Medicine, Washington University, St. Louis, Missouri, USA; Institute of Clinical and Translational Sciences, School of Medicine, Washington University, St. Louis, Missouri, USA; Division of General Internal Medicine, School of Medicine, Washington University, St. Louis, Missouri, USA; Pritzker School of Medicine, University of Chicago, Chicago, Illinois, USA

**Keywords:** Rural Health, Heart Failure, Young Adults, Hospitalization Outcomes, Access to Care

## Abstract

**Background:** Hospitalizations and mortality due to heart failure (HF) are rising in rural areas. However, inpatient outcomes for young adults with HF are not well understood. We aimed to compare in-hospital mortality, advanced procedure utilization, length of stay, and total charges among rural and urban HF patients ages 18-45.

**Methods:** We analyzed hospitalizations from the National Inpatient Sample (2016-2022), categorizing discharges as rural (National Center for Health Statistics [NCHS] 5-6), small and medium metropolitan (NCHS 3-4), and urban (NCHS 1-2). Generalized estimating equations were used to model outcomes and adjust for demographics, comorbidities, and hospital characteristics. Outcomes are reported as adjusted rate (aIRRs) or risk ratios (aRRs) with 95% confidence intervals.

**Results:** Among 79,258 HF hospitalizations among young adults, 45,075 and 10,722 were for patients from urban and rural areas, respectively. Rural patients had higher rates of in-hospital mortality (1.6% vs. 1.2%; aIRR = 1.28, 95% CI = 1.05, 1.56, p = 0.043), advanced cardiac procedure utilization (15.0% vs. 14.8%; aIRR = 1.19, 95% CI = 1.11, 1.28, p < 0.001), and longer hospital stays (aIRR = 1.10, 95% CI = 1.05, 1.14, p = 0.003). Small and medium metropolitan residents had similar outcomes to urban residents. In interaction analyses, the association between rural-urban residence and mortality differed by race (p_int_ = 0.003) and payer type (p_int_ < 0.001).

**Conclusions:** Young adults in rural areas may be prone to poor outcomes following hospitalization for HF. Strategies to identify rural adults at risk for HF and provide affordable and timely care may improve disparities.

## Introduction

Heart failure (HF) accounts for 45% of cardiovascular death in the United States,^1^ posing a substantial public health burden. Although traditionally considered a disease of older adults, the prevalence of HF among individuals ages 18-45 has increased steadily since 2012.^2^ Given that HF hospitalization is associated with increased mortality risk and substantial health care costs, the rising burden of HF among younger adults has important implications for health care utilization and spending.^2–4^

Rural residence is associated with higher risk of HF and related mortality, likely reflecting reduced access to preventive and specialty care, socioeconomic disadvantage, and structural barriers to health care.^5,6^ Rural populations experience elevated HF mortality across age groups, with particularly pronounced disparities among adults ages 35-64.^2,7^ Prior studies have also demonstrated lower utilization of advanced cardiac therapies such as cardiac resynchronization therapy (CRT) and higher rates of mortality following hospitalization for rural populations.^8^ However, findings pertaining to in-hospital outcomes for young patients from rural vs. urban areas are unknown.

Payer type, race, and hospital location may further influence the relationship between rural residence and HF hospitalization outcomes among younger adults. For example, Medicaid-insured hospitalizations are associated with decreased utilization of advanced cardiac procedures when compared to privately insured hospitalizations.^9,10^ As younger adults are more likely to be insured by Medicaid or remain uninsured than older adults,^11^ insurance coverage may play an important role in access to advanced HF care in rural settings. Racial differences in HF outcomes have also been reported, and despite having lower odds of in-hospital mortality, non-Hispanic Black and Hispanic individuals exhibit lower utilization of advanced HF therapies compared with non-Hispanic White individuals.^12^ Given that rural populations are predominantly non-Hispanic White, race may influence hospitalization outcomes.^13^ Lastly, hospitals in rural areas are more likely to lack advanced technologies and specialty care that may be needed for HF hospitalizations, and those unable to be transferred to urban hospitals or academic medical centers may face worse outcomes as a result.^14,15^

To our knowledge, no prior study has directly compared in-hospital outcomes among rural and urban individuals ages 18-45. As young adults in rural settings may be uniquely vulnerable to HF mortality, identifying differences in hospitalization outcomes is essential to inform targeted clinical and policy interventions. Accordingly, we compared in-hospital mortality, utilization of advanced cardiac procedures, length of stay, and total charges, among adults under the age of 45 hospitalized due to HF in rural, small and medium metropolitan, and urban settings.

## Methods

### Study Population

We conducted a cross-sectional analysis based on hospital admissions and associated patient characteristics, mortality, and advanced procedure utilization information provided by the Healthcare Cost and Utilization Project’s (HCUP) National Inpatient Sample (NIS).^16^ The NIS is the largest publicly available all-payer inpatient health database in the United States, comprised of a 20% sample of hospitalizations from participating hospitals; importantly, these hospitals comprise 97% of all hospital discharges nationwide. Importantly, the size of the dataset and year-to-year standardization provide country-wide generalizability and allow for aggregated analysis across years.^16^ All exposure and outcome variables, as well as covariates, were captured from the NIS. We selected the years 2016 through 2022 for analysis, as this period represents the most recent year of National Inpatient Sample data available at the time of the study that use the most updated diagnosis codes (International Classification of Diseases, Tenth Revision, Clinical Modification [ICD-10-CM]).^17^

We restricted our study population to hospitalizations with heart failure in the primary diagnosis field (ICD-10-CM diagnoses code prefixes I11.0, I13.0, I13.2, I50.1, I50.20, I50.21, I50.22, I50.23, I50.30, I50.31, I50.32, I50.33, I50.40, I50.41, I50.42, I50.43, I50.9) or with cardiogenic shock (ICD-10-CM diagnosis code prefix R570) as a primary diagnosis with a secondary diagnosis of heart failure. National Inpatient Sample data lack unique patient identifiers; thus, multiple hospitalizations of the same patient may be included. Hospitalizations in which the patient was transferred out of the hospital, outside of the ages of 18 to 45 at the time of discharge, or missing information on rural-urban residence, total charges, length of stay, payer type, sex, and race were excluded from our analysis. Inclusion and exclusion criteria are elaborated in Figure 1. This study was deemed to not be human subjects research by the Washington University Institutional Review Board (202607146).

**Figure 1:**
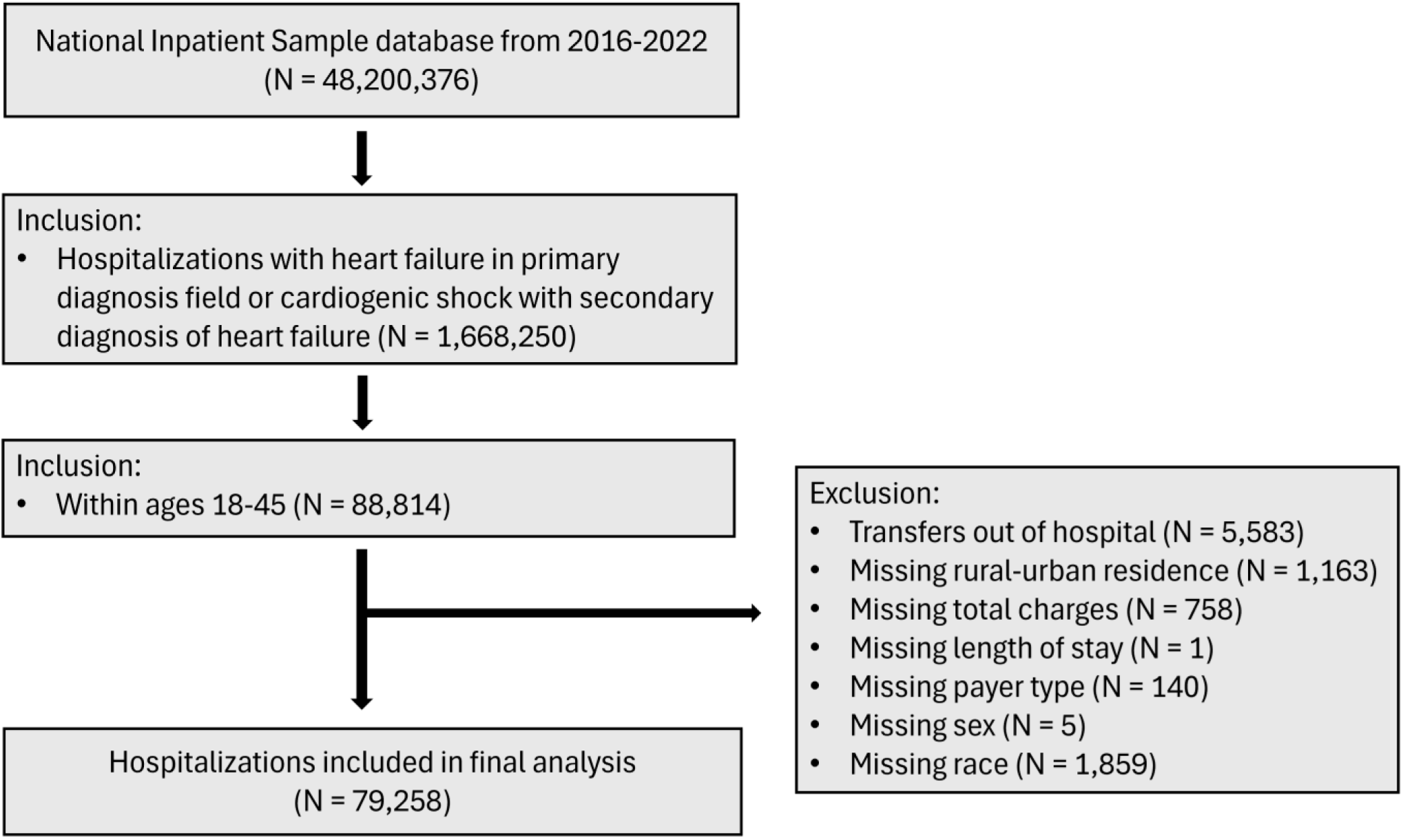
Inclusion/Exclusion Criteria. This figure presents the methodology used to select the study population. Briefly, we ascertained all hospital discharges listed in the National Inpatient Sample (NIS) from 2016-2022 with a primary diagnosis of heart failure or cardiogenic shock with a secondary diagnosis of heart failure between the ages of 18-45. We then excluded individuals who were transferred out of the hospital and those missing key demographic features.

### Exposure

Our exposure of interest was rural-urban patient residence, which was categorized based on the 2013 National Center for Health Statistics (NCHS) Urban-Rural Classification Scheme for Counties and is included in the NIS database. The NCHS classification scheme is unique in that it distinguishes between central and fringe counties within large urban areas and subdivides smaller urban counties.^18^ Since the NCHS classification uses six distinct levels, we considered multiple categorization schemes for our analysis, prioritizing interpretability while preserving meaningful distinctions between categories.^19^ We created a 3-level indicator that best reflects the observed patterning of outcomes and covariates within our dataset, grouping NCHS levels 1-2 as urban, 3-4 as small and medium metropolitan, and 5-6 as rural.^20^

### Outcomes

Our primary outcome was in-hospital mortality. Secondary outcomes include the administration of advanced cardiovascular procedures, length of stay, and total charges. Rhythm management devices, including implantable cardioverter defibrillators (ICDs), cardiac resynchronization therapy (CRT), and permanent pacemakers (PPMs), were identified using ICD-10-CM codes with prefixes 0JH6 and 0JH7. Mechanical circulatory support device procedures were captured using ICD-10-CM procedure codes 5A02210, 5A02110, and codes with the prefix 5A1522, corresponding to intra-aortic balloon pumps (IABPs), percutaneous left ventricular assist devices (pLVADs), and extracorporeal membrane oxygenation (ECMO). Coronary angiography (CA) was identified using ICD-10-CM code prefix B21. Left ventricular assist devices (LVADs) were captured with ICD-10-CM code prefix 02HA, and heart transplantation was identified using ICD-10-CM code prefix 02YA.^17^

### Covariates

Covariates used in the present study include patient and hospital characteristics, as well as relevant comorbidities. We used a 2-level indicator for age (18-34 and 35-45). Additional patient characteristics include sex, race/ethnicity (non-Hispanic White, non-Hispanic Black, Hispanic, Asian or Pacific Islander, Native American, and Other), payer type (Medicare, Medicaid, private insurance, self-pay, no charge, and Other), hospital bed size (small, medium, and large), hospital location (rural, urban nonteaching, and urban teaching), and hospital region (Northeast, Midwest, South, and West). Female sex, non-Hispanic White race, private insurance, large bed size (400+ beds), urban teaching hospitals, and the South region were used as reference categories in subsequent analysis due to large representation in our dataset and clinical relevance. Relevant comorbidities identified using the Elixhauser Comorbidity Index included hypertension, arrhythmias, renal failure, diabetes, chronic obstructive pulmonary disease (COPD), fluid and electrolyte disorders, valvular disorders, obesity, pulmonary circulatory disease, peripheral vascular disease, hypothyroidism, depression, deficiency anemia, coagulopathy, liver failure, alcohol abuse, and drug abuse.^21^

### Statistical Analysis

We first generated descriptive statistics for our final study population. We then calculated crude outcomes of interest by rural-urban residence. For categorical characteristics, we present percentages of hospitalizations and standardized mean differences. We chose to present both mean length of stay with standard deviation (SD) and median total charges with p-values from the Kruskal-Wallis test. For categorical outcomes, we present percentages of hospitalizations and p-values generated from chi-squared testing, while for continuous outcomes, we present summary statistics and p-values generated from the Kruskal-Wallis test. Sample sizes are reported as unweighted counts, whereas all adjusted analyses incorporate NIS-provided discharge weights, which are used to adjust unweighted discharge records so they represent national estimates.

We used generalized estimating equations (GEE) to create 1) a quasi-Poisson regression to estimate the effect of rural-urban residence on all countable outcomes (in-hospital mortality, utilization of advanced cardiac procedures, length of stay), and 2) a log-linked regression with the gamma distribution for total charges. Prior to modelling, we aggregated hospitalizations by hospital-year, sex, race, payer type, hospital characteristics, and relevant comorbidities. We adjusted for all covariates, clustered on hospital ID with an exchangeable correlation structure within each year to account for patterns in outcomes throughout the study period, and weighted results based on HCUP-provided weights. We also examined the interactions between rural-urban residence and payer type, race, and hospital location. Due to the low outcome count among certain payer (self-pay, no charge) and race (Asian, Native American) categories within rural areas, we collapsed these smaller categories into “Other” categories when assessing interactions. Lastly, we conducted sensitivity analysis for hospital transfers by modeling outcomes excluding patients transferred into inpatient settings from a different acute care facility. Results are reported as adjusted incidence rate ratios (aIRRs) for count outcomes and rate ratios (aRRs) for continuous outcomes (i.e., total charges), each with 95% confidence intervals. To account for multiple hypotheses, we applied a false discovery rate adjustment for our primary and secondary outcomes of interest in both adjusted and interaction models. Since specific advanced procedures are subsets of the larger advanced procedure outcome, we considered analyses of these subgroups to be exploratory and hypothesis-generating; therefore, we did not apply formal adjustment for multiplicity to these outcomes.

We used Python 3.13 for all analyses, including plotting, model generation, and statistical analysis, and utilized the Elixhauser Comorbidity Index package in SAS.^21,22^

## Results

### Patient Characteristics

Our unweighted analytic sample included 79,258 hospitalizations due to HF among adults between the ages of 18 and 45 between January 2016 and December 2022. Within our study population, 45,075 (56.9%) of hospitalizations were from urban residents, while 23,461 (29.6%) and 10,722 (13.5%) were attributed to small and medium metropolitan and rural residents, respectively. When comparing rural and urban residents, the proportion of rural patients were ages 35 years and older (76.6% vs. 74.2%, SMD = 0.056) and female patients (39.0% vs. 36.0%, SMD = 0.062) were similar between groups. Rural patients were more predominately non-Hispanic White (52.7% vs. 24.1%), Native American (3.9% vs. 0.4%, SMD = 1.253), and enrolled in Medicare (22.6% vs. 18.5%) or self-pay (15.7% vs. 12.9%, SMD = 0.232). Furthermore, rural residents showed fewer reported diagnoses of alcohol abuse, arrhythmia, anemia, fluid and electrolyte disorders, hypertension, obesity, pulmonary circulation disease, and renal failure, as well as higher incidence of reported chronic obstructive pulmonary disease (COPD), depression, drug abuse, hypothyroidism, and peripheral vascular disease.

A greater proportion of discharges among rural residents came from larger hospitals (65.7% vs. 52.4%, SMD = 0.715), rural hospitals (45.1% vs. 0.3%, SMD = 1.552), and hospitals in the South (67.2% vs. 42.5%, SMD = 0.606, Table 1). Small and medium metropolitan residents were demographically similar in age distribution to urban patients but had a higher proportion of non-Hispanic White patients and lower proportions of non-Hispanic Black and Hispanic patients (SMD = 1.253). Clinically, they demonstrated lower prevalence of alcohol abuse, arrhythmia, anemia, fluid and electrolyte disorders, hypertension, pulmonary circulation disease, and renal failure, but higher rates of chronic obstructive pulmonary disease, depression, drug abuse, and hypothyroidism compared with urban residents; most hospitalizations of small and medium metropolitan patients occurred in urban teaching hospitals and in the South.

**Table 1:** Unweighted Characteristics of Young Adults (18-45) Hospitalized Due to Heart Failure, 2016-2022 (N = 79,258)

| Characteristics (n,%) |  | Urban (n = 45,075) | Small and Medium Metropolitan (n = 23,461) | Rural (n = 10,722) | Standardized Mean Differences (SMD) |
| --- | --- | --- | --- | --- | --- |
| Age Category | 18-34 | 11625 (25.8) | 5955 (25.3) | 2512 (23.4) | 0.056 |
| Sex | Female | 16229 (36.0) | 8818 (37.6) | 4176 (39.0) | 0.062 |
| Race | White | 10887 (24.1) | 9135 (38.9) | 5646 (52.7) | 1.253 |
|  | Black | 23417 (52.0) | 10025 (42.7) | 3791 (35.4) |  |
|  | Hispanic | 7503 (16.7) | 2842 (12.1) | 585 (5.46) |  |
|  | Asian | 1476 (3.27) | 700 (2.98) | 120 (1.11) |  |
|  | Native American | 199 (0.44) | 336 (1.43) | 414 (3.86) |  |
|  | Other | 1593 (3.53) | 423 (1.80) | 166 (1.08) |  |
| Payer Type | Medicare | 8320 (18.5) | 4768 (20.3) | 2428 (22.6) | 0.232 |
|  | Medicaid | 19937 (44.2) | 10433 (44.5) | 4208 (39.2) |  |
|  | Private insurance | 9717 (21.6) | 4566 (19.5) | 2057 (19.2) |  |
|  | Self-pay | 5839 (12.9) | 2966 (12.6) | 1679 (15.7) |  |
|  | No charge | 389 (0.86) | 158 (0.77) | 68 (0.63) |  |
|  | Other | 873 (1.94) | 570 (2.43) | 282 (2.63) |  |
| Comorbidities | Hypertension | 38207 (84.8) | 19561 (83.4) | 8730 (81.4) | 0.091 |
|  | Renal failure | 24322 (54.0) | 11939 (50.9) | 5195 (48.4) | 0.112 |
|  | Obesity | 20391 (45.2) | 10448 (44.5) | 4708 (43.9) | 0.026 |
|  | Fluid and Electrolyte Disorders | 18107 (40.2) | 9330 (39.8) | 4166 (38.8) | 0.029 |
|  | Diabetes | 16006 (35.5) | 8589 (36.6) | 3854 (35.9) | 0.023 |
|  | Chronic Obstructive Pulmonary Disease | 13265 (29.4) | 7910 (33.7) | 3572 (33.3) | 0.093 |
|  | Arrhythmias | 10299 (22.8) | 5132 (21.9) | 2220 (20.7) | 0.051 |
|  | Pulmonary circulation disease | 9758 (21.6) | 4824 (20.6) | 2169 (20.2) | 0.034 |
|  | Drug abuse | 9294 (20.6) | 5523 (23.5) | 2279 (21.3) | 0.070 |
|  | Valvular disease | 7841 (17.4) | 4220 (18.0) | 1833 (17.1) | 0.024 |
|  | Depression | 5837 (12.9) | 3430 (14.6) | 1537 (14.3) | 0.049 |
|  | Liver disease | 4700 (10.4) | 2499 (10.6) | 1131 (10.5) | 0.007 |
|  | Deficiency Anemia | 4018 (8.91) | 1834 (7.82) | 715 (6.67) | 0.082 |
|  | Alcohol Abuse | 3571 (7.92) | 1764 (7.52) | 771 (7.19) | 0.026 |
|  | Hypothyroidism | 2847 (6.32) | 1858 (7.92) | 958 (8.93) | 0.098 |
|  | Coagulopathy | 2769 (6.14) | 1423 (6.07) | 639 (5.96) | 0.004 |
|  | Peripheral vascular disease | 1157 (2.56) | 614 (2.62) | 334 (3.12) | 0.030 |
| **Bed Size** | Small | 8728 (19.4) | 4721 (20.1) | 1346 (12.5) | 0.715 |
|  | Medium | 12714 (28.2) | 6337 (27.1) | 2330 (21.7) |  |
|  | Large | 23633 (52.4) | 12403 (52.9) | 7046 (65.7) |  |
| **Hospital Location** | Rural | 122 (0.27) | 289 (1.23) | 4832 (45.1) | 1.552 |
|  | Urban Nonteaching | 7979 (17.7) | 5273 (22.5) | 917 (8.55) |  |
|  | Urban Teaching | 36974 (82.0) | 17899 (76.3) | 4973 (46.4) |  |
| **Hospital Region** | Northeast | 6476 (14.4) | 2022 (8.62) | 467 (19.5) | 0.606 |
|  | Midwest | 8918 (19.8) | 4035 (17.2) | 2104 (19.6) |  |
|  | South | 19173 (42.5) | 12839 (54.7) | 7201 (67.2) |  |
|  | West | 10508 (23.3) | 4565 (19.5) | 950 (8.86) |  |

### Unadjusted Outcomes

Figure 2 shows unadjusted primary and secondary outcomes following hospitalization due to HF by rural-urban residence. Rural patients had higher rates of in-hospital mortality compared to their small and medium metropolitan and urban counterparts (1.6% vs. 1.3% vs. 1.2%, p = 0.001). Overall utilization of advanced cardiac procedures was similar in all groups (15.0% vs. 14.7% vs. 14.8%, p = 0.875), though small and medium metropolitan and rural residents show a slight reduction in rhythm management device procedures (2.9% vs. 2.9% vs. 3.2%, p = 0.041). Mean length of stay did not differ by urban-rural location in our unadjusted results (5.2 days vs. 5.1 vs. 5.3, p = 0.101), though total charges were significantly lower for rural residents ($27,459.5 vs. $32,036.0 vs. $38,422.0, p <0.001). Notably, a smaller proportion of rural residents left against medical advice (5.6% vs. 6.8% vs. 7.2%, p <0.001, Supplemental Table 1).

**Figure 2:**
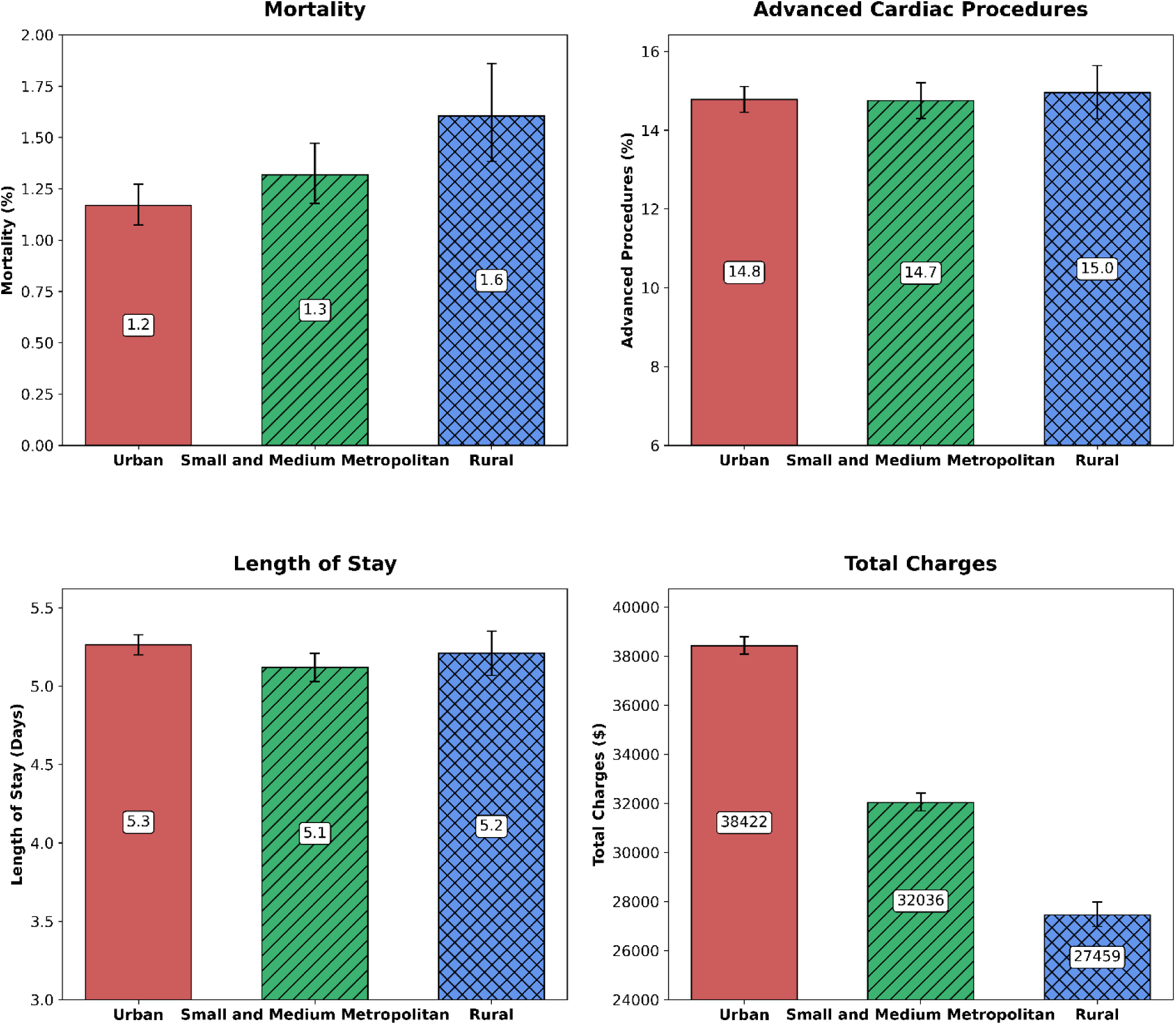
Unadjusted Outcomes for Young Adults (18-45) During Hospitalization Due to Heart Failure by Rural-Urban Residence, 2016-2022. We present unadjusted primary (mortality) and secondary (advanced cardiac procedures, length of stay, and total charges) following inpatient hospitalization due to heart failure, stratified by rural-urban residence. Hospitalizations of rural residents show increased rates of mortality and lower total charges, with similar advanced procedure utilization frequency and lengths of stay to both urban and small and medium metropolitan subgroups.

### Adjusted Outcomes

Adjusting for sex, race, payer type, patient characteristics, and comorbidities, rural, but not small and medium metropolitan residents, had higher rates of in-hospital mortality (aIRR rural = 1.28, 95% CI = 1.05, 1.56, p = 0.043; aIRR small and medium metropolitan = 1.04, 95% CI = 0.90, 1.20, p = 0.584) and advanced cardiac procedure utilization (aIRR rural = 1.19, 95% CI = 1.11, 1.28, p < 0.001; aIRR small and medium metropolitan = 1.01, 95% CI = 0.97, 1.06, p = 0.584) compared to urban patients. Concerning exploratory findings for specific procedures, rural hospitalizations had higher rates of rhythm management device (aIRR = 1.29, 95% CI = 1.11, 1.49) and coronary angiography procedures (aIRR = 1.18, 95% CI = 1.10, 1.28), while hospitalization for people from small and medium metropolitan counties had lower rates of mechanical circulatory support procedures (aIRR = 0.84, 95% CI = 0.71, 0.99). Lengths of stay for rural residents were significantly longer (aIRR rural = 1.10, 95% CI = 1.05, 1.14, p = 0.003) and following adjustment, total charges per hospitalization were slightly higher among rural residents (aRR rural = 1.05, 95% CI = 1.00, 1.11, p = 0.144, Table 2). Outcomes did not differ when excluding patients who were transferred in from other acute care facilities (Supplemental Table 3).

**Table 2:** Adjusted In-Hospital Outcomes for Hospitalizations Due to Heart Failure by Rural-Urban Residence, 2016-2022.

| **Rural-Urban Residence** | **Small and Medium Metropolitan**<br>**(n = 23,461)** |  | **Rural**<br>**(n = 10,722)** |  |
| --- | --- | --- | --- | --- |
| **Outcome (Ref = Urban)** | **Effect<sup>a</sup> (95% CI)** | ***p*** | **Effect (95% CI)** | ***p*** |
| Mortality | 1.04 (0.90, 1.20) | 0.584 | 1.28 (1.05, 1.56) | 0.043 |
| Received Advanced Cardiac Procedures | 1.01 (0.97, 1.06) | 0.584 | 1.19 (1.11, 1.28) | <0.001 |
| ICD/CRT/PPM | 0.98 (0.89, 1.08) |  | 1.28 (1.11, 1.49) |  |
| IABP/pLVAD/ECMO | 0.84 (0.71, 0.99) |  | 1.20 (0.96, 1.50) |  |
| Coronary Angiography | 1.04 (0.99, 1.10) |  | 1.18 (1.10, 1.28) |  |
| Left Ventricular Assist Device | 1.02 (0.83, 1.25) |  | 1.24 (0.94, 1.64) |  |
| Heart Transplant | 0.90 (0.70, 1.15) |  | 0.85 (0.59, 1.23) |  |
| Length of Stay | 0.99 (0.96, 1.01) | 0.387 | 1.10 (1.05, 1.14) | 0.003 |
| Total Charges<sup>a</sup> | 0.98 (0.94, 1.02) | 0.387 | 1.05 (1.00, 1.11) | 0.144 |

Quasi-Poisson and Gamma log-link Generalized Estimating Equations (GEEs) aggregated by hospital-year, sex, race, payer type, hospital bed size, region, and location, and comorbidities (hypertension, arrhythmias, renal failure, diabetes, chronic obstructive pulmonary disease [COPD], fluid and electrolyte disorders, valvular disorders, obesity, pulmonary circulatory disease, peripheral vascular disease, hypothyroidism, depression, deficiency anemia, coagulopathy, liver failure, alcohol abuse, and drug abuse), and adjusted for patient and hospital characteristics. Urban counties are used as a reference. All models account for clustering by hospital-year and HCUP-provided weights. Sample sizes are reported as unweighted.
<sup>a</sup>The measure of association for count outcomes (mortality, advanced procedures, and length of stay) is an adjusted incidence rate ratio (aIRR), and the measure of association for continuous outcomes (total charges) is an adjusted rate ratio (aRR).

### Race

We assessed interactions between rural–urban residence and race across in-hospital outcomes. Significant interactions were identified for in-hospital mortality (p_int_ = 0.003), though not for advanced cardiac procedures, length of stay, or total charges (Supplemental Table 2). Mortality was higher, though non-significantly, for rural residents of non-Hispanic Black, Hispanic, and other races, compared to non-Hispanic White patients in urban areas.

### Payer Type

We found significant interactions between rural-urban residence and payer type pertaining to mortality and advanced cardiac procedures (p_int_ < 0.05, Supplemental Table 2). Rural residents on Medicare, Medicaid, or other insurance types did not show significantly higher rates of mortality, length of stay, or total charges when compared to urban residents utilizing private insurance, though patients not insured by Medicare, Medicaid, or private insurance show elevated rates of advanced procedure utilization (aIRR = 1.28, 95% CI = 1.09, 1.51, Figures 3A-D, Supplemental Table 2).

**Figures 3A-D.**
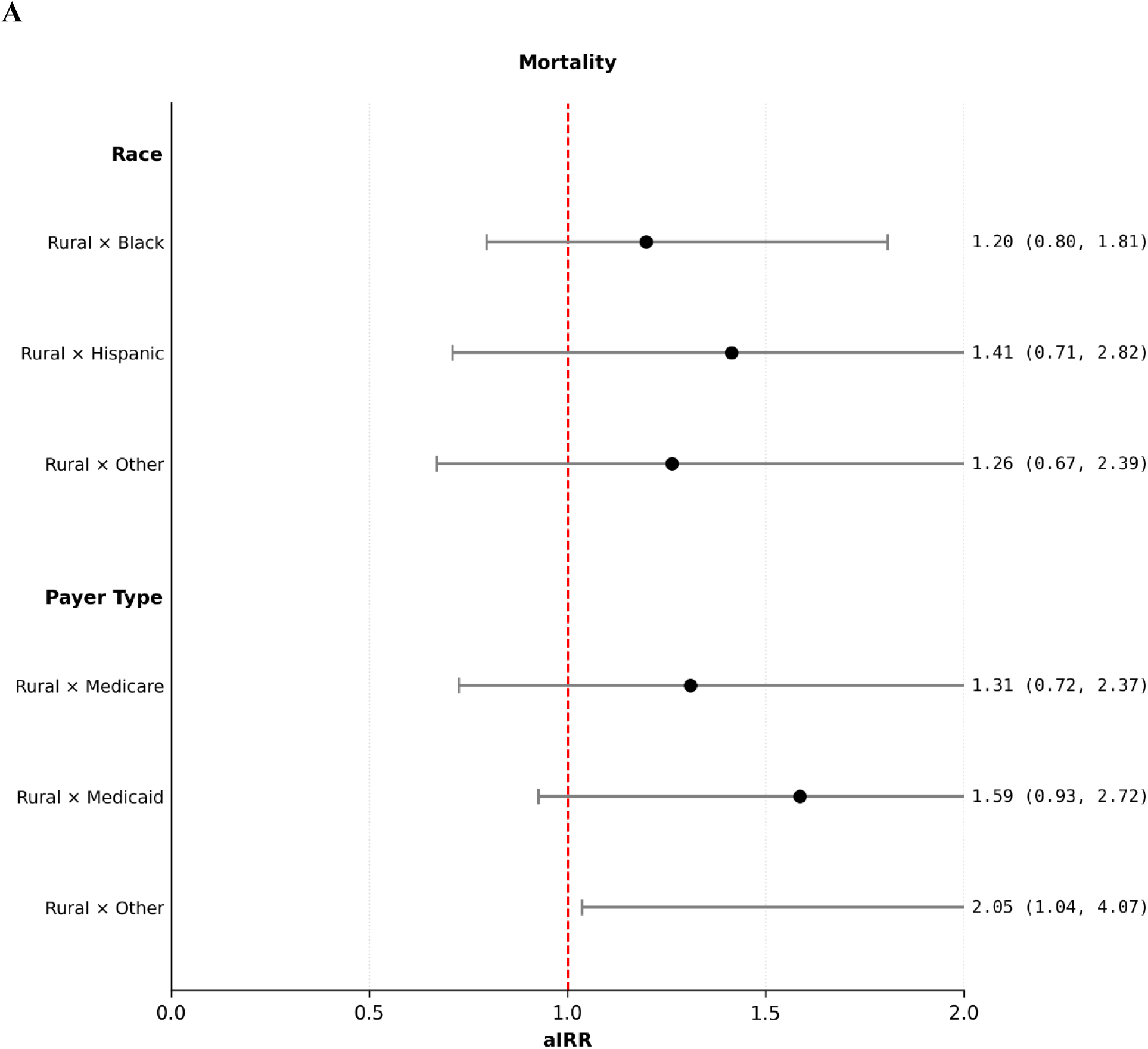
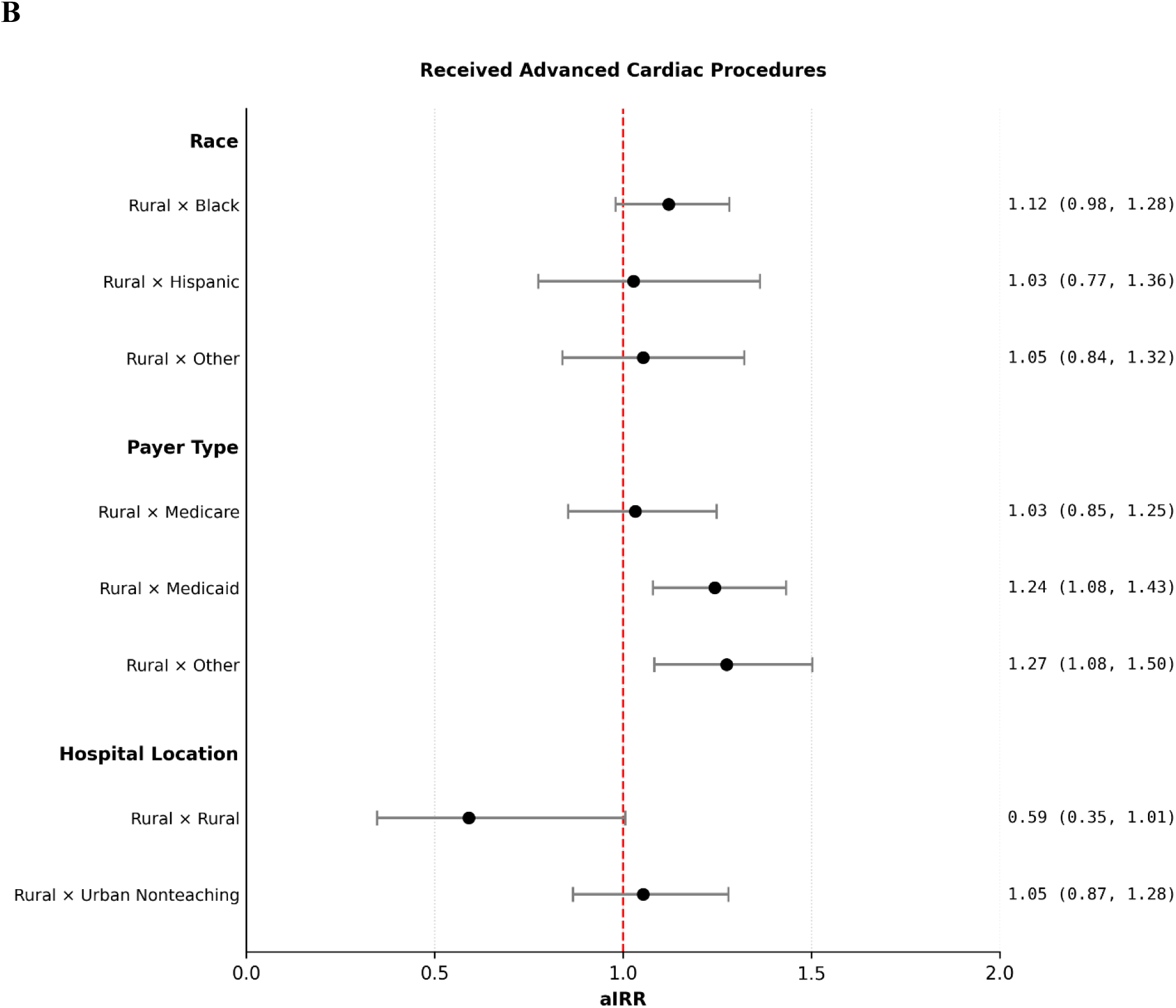
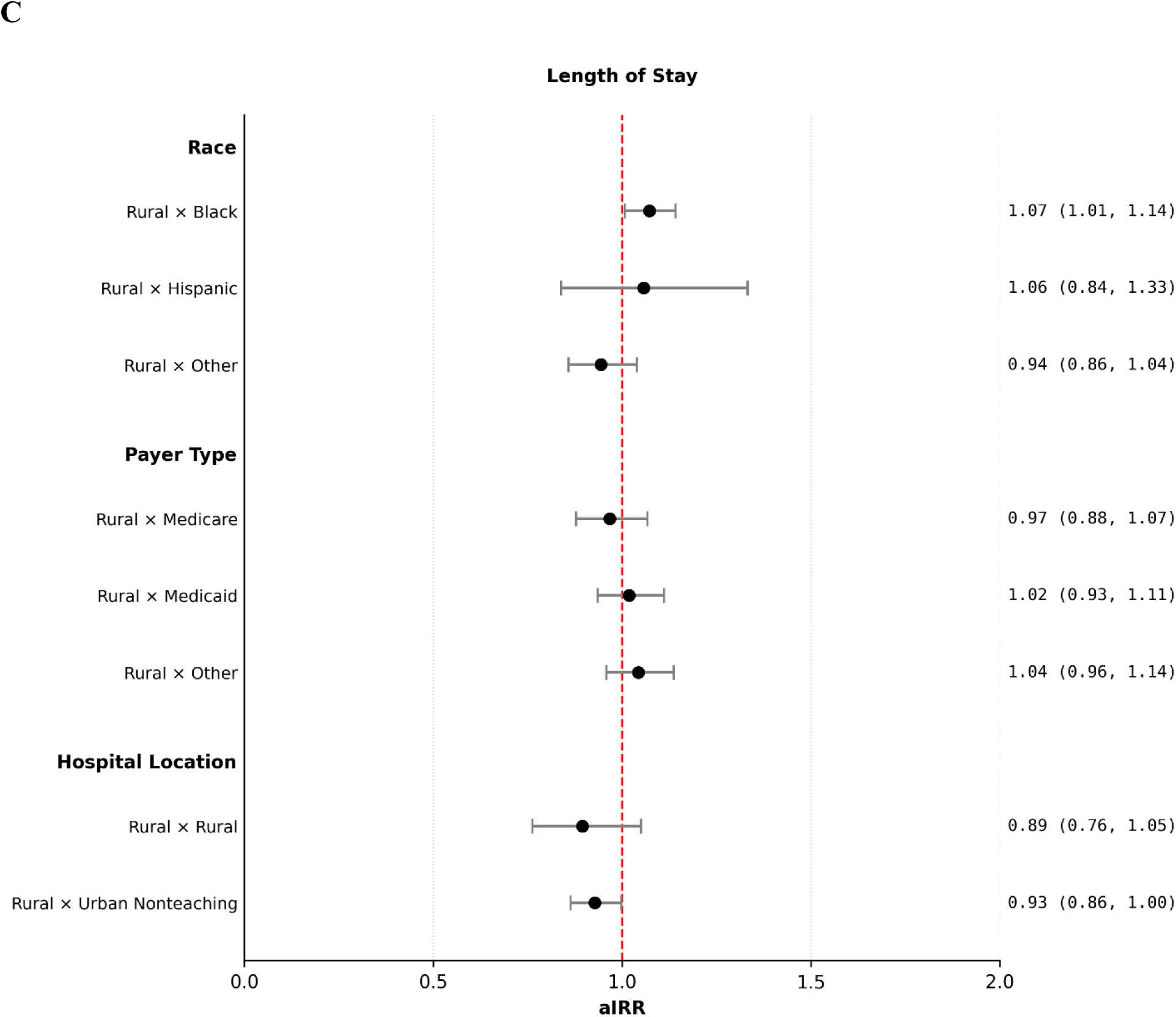
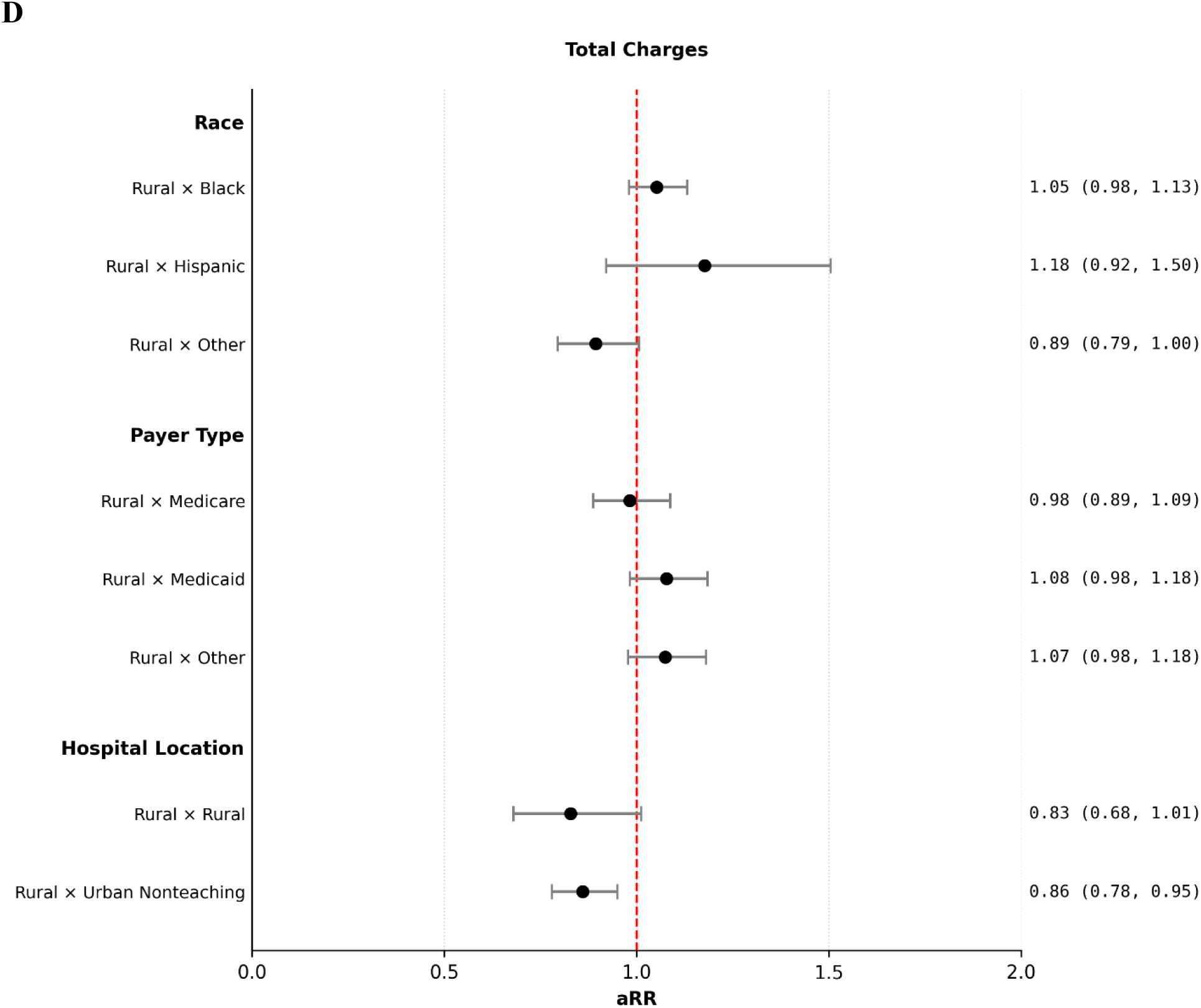
Adjusted Interaction Effects of Rural-Urban Residence on Outcomes Following Hospitalizaiton of Young Adults (18-45) Due to Heart Failure (Unweighted N = 79,258) Quasi-Poisson (A: mortality; B: received advanced cardiac procedures; C: length of stay) and Gamma log-link (D: total charges) generalized estimating equations (GEEs) evaluating interactions between rural-urban residence and race, payer type, and hospital location. As mortality outcomes were rare for rural patients discharged from rural hospitals, these outcomes were omitted from figure 3A. The data shows strong heterogeneity in outcomes when analyzing interactions, suggesting that outcomes are driven by a combination of factors and may vary greatly between rural-urban subgroups.

### Hospital Location

We also examined interactions between patient rural–urban residence and hospital location. While there were no significant interactions between rural-urban residence and hospital location for mortality (p_int_ = 0.912) or advanced procedure utilization (p_int_ = 0.333), we observed significant interactions for length of stay (p_int_ = 0.038) and total charges (p_int_ < 0.001, Supplemental Table 2). Rural patients discharged from urban nonteaching hospitals showed slightly shorter hospital stays (aIRR = 0.93, 95% CI = 0.87, 1.00) and significantly lower total charges than urban patients in urban teaching hospitals (aRR = 0.87, 95% CI = 0.79, 0.96, Figures 3B-D, Supplemental Table 2).

## Discussion

In this analysis of HF hospitalizations among young adults (18-45 years), rural residence was associated with higher in-hospital mortality, higher utilization of advanced cardiac procedures, and longer hospital stays. For discharges of residents from small and medium metropolitan areas, hospitalization outcomes were similar to those of urban residents. The association between rural-urban residence and selected outcomes varied by race, payer type, and hospital location, although these interactions were limited to specific outcomes. Overall, these findings highlight persistent rural-urban differences in hospital outcomes among young adults with HF.

Although prior data on young adults with HF residing in rural areas are limited, our findings align with earlier reports demonstrating higher mortality among residents of more rural counties, including those that include younger populations.^6–8,23^ Differences in rural classification schemes, covariate adjustment, and age group definitions across studies may account for some variation in estimates; however, the overall pattern of elevated mortality in the most rural areas of the United States remains consistent. Our study extends prior work by focusing specifically on hospitalized young adults and examining whether rural-urban differences varied by payer type, race and ethnicity, and hospital location.

The interaction analyses suggested that rural-urban differences in selected outcomes varied by race, payer type, and hospital location. However, these interactions were limited to specific outcomes. Thus, these findings should be interpreted as evidence of potential heterogeneity in rural-urban differences rather than as evidence that these factors independently explain observed differences.

Rural patients were more likely to undergo advanced cardiac procedures, including rhythm management devices and coronary angiography, raising the possibility that differences in disease severity or presentation contributed to the observed rural-urban differences. While previous research has shown that rural patients may be hospitalized at a lower severity threshold, most subjects in prior studies were above the age of 65. In these studies, researchers hypothesized that rural patients may be hospitalized at a lower severity threshold due to anticipated access issues, and it is unclear whether this phenomenon is occurring in younger populations.^24^ Future studies incorporating measures of HF severity at admission, including laboratory results and hemodynamics, are needed to clarify the role of delayed or inadequate outpatient management.

Rural residents had longer hospital stays than urban residents, coinciding with previous findings. The longer length of stay among rural residents may reflect differences in care coordination, discharge planning, access to post-acute services, payer type, or availability of outpatient follow-up after discharge.^25–27^

Despite higher mortality and use of advanced procedures, unadjusted charges were lower for rural versus urban patients, with no difference between urban and small and medium metropolitan residents. Because hospital charges reflect differences in hospital systems, resource utilization, reimbursement, and patient and hospital characteristics, these findings are difficult to interpret as a direct measure of the quality or efficiency of care.

Together, these findings suggest that young adults in the most rural communities may require more aggressive, intensive care upon entering the hospital, potentially reflecting limited access to outpatient care including specialty cardiovascular care, transportation, and insurance coverage. Results underscore the need for targeted strategies to improve access to preventive and specialty HF care in rural communities and to address intersecting socioeconomic and racial disparities in this growing population. Potential interventions include telecardiology programs to engage young adults in care, investment in diagnostic and treatment resources for young adults at rural hospitals, and expanded insurance coverage and financial support to reduce barriers to access. Additional approaches could involve coordinated transportation for appointments, community-based education on early HF risk, and integration of risk stratification tools into clinical practice to identify and manage high-risk young adults prior to development of advanced disease.

This study has several important limitations. First, the NIS relies on billing codes for diagnoses and procedures, which introduces the potential for misclassification and miscoding. As the NIS dataset includes inpatient discharge-level data only, analyses were conducted at the hospitalization level rather than the patient level, and we could not track transfers between hospitals. However, our exclusion of transfers-out should avoid inappropriately undercounting interventions for patients who initially presented to a small rural hospital and were transferred for definitive care. We also could not assess disease duration or prior procedures, or longitudinal outcomes after discharge, and lacked clinical details including ejection fraction, laboratory values, medication use, and outpatient management history. Total charges were not adjusted for inflation and therefore may reflect changes in the purchasing power of the dollar over the study period. Rural-urban residence was defined using the NCHS county-level classification, which does not capture within-county heterogeneity in rurality. Similarly, hospital location is categorized broadly as rural or urban, with urban hospitals further subdivided into teaching and nonteaching institutions. This structure does not account for gradients of rurality within these categories and may obscure meaningful geographic variation. Sparse data in certain subgroups, including self-pay or no-charge encounters, Asian and Native American patients, and urban nonteaching and rural hospitals, combined with rare outcomes, limited statistical power for interaction testing. Analyses of individual advanced procedures were not adjusted for multiplicity and are exploratory and hypothesis-generating. Finally, relevant individual circumstances, including income and education, and behaviors, such as diet and exercise, were not available for this analysis.

## Conclusion

Young adults living in rural areas of the United States experience elevated rates of HF-related in-hospital mortality and advanced procedure utilization, as well as longer hospital stays, compared to their urban counterparts. Insurance coverage, race and ethnicity, and hospital location may contribute to these outcomes. Findings warrant targeted clinical and community-level interventions to prevent development of severe HF prior to hospitalization.

## Clinical Perspective

- Among younger adults, rural residence was associated with higher in-hospital mortality, increased utilization of advanced cardiac procedures, and longer hospital stays. These disparities may particularly impact patients with public health insurance and racial or ethnic minorities.
- Rural health systems with disadvantaged patients) should consider clinical interventions to identify young patients at risk of HF and engage them in preventive and specialty care.
- Additional information on disease severity (ejection fraction and laboratory measures), as well as long-term follow-up post-hospitalization, are needed to identify underlying mechanisms behind poor in-hospital outcomes for young, rural patients hospitalized due to HF.

## Data Availability

The data used in this study are publicly available from the Healthcare Cost and Utilization Project (HCUP) National Inpatient Sample (NIS), maintained by the Agency for Healthcare Research and Quality (AHRQ). Access to the NIS requires registration and completion of the HCUP Data Use Agreement. The data are available from HCUP upon completion of the required data-use procedures.

https://hcup-us.ahrq.gov/nisoverview.jsp

